# Poor Hemorrhagic Stroke Outcomes During the COVID-19 Pandemic Are Driven by Socioeconomic Disparities: Analysis of Nationally Representative Data

**DOI:** 10.1101/2023.07.07.23292399

**Authors:** Abdulaziz T. Bako, Thomas Potter, Alan Pan, Karim A. Borei, Taya Prince, Gavin Britz, Farhaan S. Vahidy

**Affiliations:** Department of Neurosurgery, Houston Methodist, Houston, TX; Center for Health Data Science and Analytics, Houston Methodist, Houston, TX; Department of Population Health Sciences, Weill Cornell Medical College, New York, NY

**Keywords:** Intracerebral Hemorrhage, Subarachnoid Hemorrhage, COVID-19, Socioeconomic Disparities, Social Determinants of Health

## Abstract

**Background:** Nationally representative data demonstrating the impact of the COVID-19 pandemic on hemorrhagic stroke outcomes are lacking.

**Methods:** In this pooled cross-sectional analysis, we used the National Inpatient Sample (2016-2020) to identify adults (>=18 years) with primary intracerebral hemorrhage (ICH) or subarachnoid hemorrhage (SAH). We fit segmented logistic regression models to evaluate the differences in the rates of in-hospital outcomes (in-hospital mortality, home discharge, and receiving neurosurgical procedures) between the pre-pandemic (January 2016-February 2020) and pandemic periods (March 2020-December 2020). We used multivariable logistic regression models to evaluate the differences in mortality between patients admitted from April to December 2020, with and without COVID-19, and those admitted during a similar period in 2019. Stratified analyses were conducted among patients residing in low and high-income zip codes and among patients with extreme loss of function (E-LoF) and those with minor to major loss of function (MM-LoF).

**Results:** Overall, 309,965 ICH patients (mean age [SD]: 68[14.8], 47% female, 56% low-income) and 112,210 SAH patients (mean age [SD]: 60.2[15.4], 62% female, 55% low-income) were analyzed. Pre-pandemic, ICH mortality was decreasing by ≈ 1 % per month (adjusted odds ratio, 95% confidence interval: 0.99, 0.99-1.00). However, during the pandemic, the overall ICH mortality rate increased by ≈ 2% per month (1.02, 1.00-1.02) and ≈ 4% per month among low-income patients (1.04, 1.01-1.07). However, there was no change in trend among high-income ICH patients during the pandemic (1.00, 0.97-1.03). Patients with comorbid COVID-19 in 2020 had significantly higher odds of mortality compared to the 2019 comparison cohort, overall (ICH: 1.83, 1.33-2.51; SAH: 2.76, 1.68-4.54), and among patients with MM-LoF (ICH: 2.15, 1.12-4.16; SAH: 5.77, 1.57-21.17). However, patients with E-LoF and comorbid COVID-19 had similar mortality rates with the 2019 cohort.

**Conclusion:** Sustained efforts are needed to address socioeconomic disparities in healthcare access, quality, and outcomes during public health emergencies.

## Introduction

The global outbreak of COVID-19, caused by the novel coronavirus SARS-CoV-2, resulted in a pandemic that disrupted healthcare, especially among vulnerable populations.^1,2^ COVID-19 infection may worsen vascular diseases by disrupting the coagulation cascade and exacerbating inflammatory responses.^3,4^ Although prior studies have shown that COVID-19 increases the risk of poor outcomes among patients with ischemic stroke,^5,6^ there is a paucity of nationally representative data on the potential impact of the COVID-19 pandemic on the trends in hemorrhagic stroke (intracerebral hemorrhage [ICH] and subarachnoid hemorrhage [SAH]) outcomes. Therefore, we used the largest publicly available all-payer inpatient healthcare database in the United States (US), the National Inpatient Sample (NIS), to evaluate the differences in the trends of hemorrhagic stroke outcomes before and after the COVID-19 pandemic.

## Methods

### Ethics Statement

Because this research utilized publicly available and de-identified data, it is considered exempt from review by the Houston Methodist Institutional Review Board. We followed the STrengthening the Reporting of OBservational studies in Epidemiology (STROBE) guidelines.^7^

### Data availability

After completing a data use agreement training, qualified researchers can obtain NIS data through the Health Care Utilization Project’s central distributor (https://www.distributor.hcup-us.ahrq.gov/).

### Study Design, Data Source, and Case Identification

NIS represents over 90% of all US hospitalizations.^8^ In this pooled cross-sectional study, we used validated International Classification of Disease Tenth Revision (ICD-10) codes to identify adults (≥ 18 years) discharged with a principal diagnosis of ICH (ICD-10 codes: I61.0-I61.6 and I61.8-I61.9) or SAH (I60) from 2016 to 2020. We excluded patients with concurrent diagnoses of head trauma and/or arteriovenous malformation, as well as patients with missing age information. Also, we excluded patients transferred to an acute care hospital to avoid double counting the same patient, as the unit of observation in the NIS database is a hospitalization encounter and not an individual patient. Among the ICH cohort, we additionally excluded patients with co-occurring diagnoses of intracranial aneurysms and brain malignancy.

Race/ethnicity was coded as non-Hispanic White (NHW), non-Hispanic Black (NHB), Asian American and Pacific Islanders (AAPI), Hispanic, and Others (including Native Americans and Others). Income status was defined according to the income quartile of the patient’s zip code, with quartiles 1 and 2 considered as low-income and quartiles 3 and 4 considered as high-income zip codes. The National Institutes of Health Stroke Scale (NIHSS) score was only available for less than one-third (20.7%) of our analysis sample; therefore, we utilized the administratively derived All Patient Refined Diagnosis Related Group (APR-DRG) severity of illness scores to assess disease severity. We further grouped patients, based on their APR-DRG severity of illness score, into those with extreme loss of function (E-LoF) and those with minor to major loss of function (MM-LoF).^9^ COVID-19 status was identified using ICD-10 code U07.1. This ICD code was released in late March of 2020 and is reserved for laboratory-confirmed cases of SARS-CoV-2.

The primary outcome is in-hospital mortality and secondary outcomes include home discharge, receiving craniotomy (for ICH cohort), and undergoing coiling or clipping (for SAH cohort).

### Statistical Analyses

Descriptive statistics were reported using means and percentages. We used a series of univariable logistic regression models to evaluate the differences in the clinical and sociodemographic characteristics of patients admitted before the official declaration of national emergency response to the COVID-19 pandemic (January 2016 - February 2020) (pre-pandemic period), and patients admitted during and after the emergency declaration (March 2020 – December 2020) (pandemic period). Furthermore, we fit a series of unadjusted and adjusted segmented logistic regression models^10^ (details in supplemental methods) to evaluate the differences in the rates (slope) of in-hospital outcomes between the pre-pandemic and pandemic periods, as crude/adjusted odds ratios (OR/aOR) and 95% confidence interval (CI). The multivariable models included adjustments for sociodemographic factors (age, sex, race/ethnicity, and insurance type), clinical factors (hypertension, diabetes, congestive heart failure, obesity, renal failure, Charlson Comorbidity Index, and APR-DRG severity of illness score), and hospital-related factors (urban/rural location of hospital, teaching status of hospital, and hospital bed size). To evaluate whether income modifies the association of the pandemic with ICH and SAH outcomes, we performed stratified analyses among patients residing in low-income and high-income zip codes.

In a second series of analyses, we used multivariable logistic regression models to evaluate the sociodemographic and clinical factors independently associated with having comorbid COVID-19 infection and ICH or SAH among a cohort of patients admitted April to December 2020. In a third and final series of analyses, we used multivariable logistic regression models to assess the differences in mortality, between ICH and SAH patients admitted from April to December 2020, with and without COVID-19, and patients admitted during a similar period in 2019. We performed stratified multivariable analyses among patients with MM-LoF and those with E-LoF. The confounding variables in all adjusted models were selected based on prior evidence demonstrating their association with hemorrhagic stroke outcomes. All analyses were conducted with 0.05 level of significance, utilizing Stata version 17.^11^.

## Results

Overall, 309,965 ICH patients (mean age [SD]: 68 [14.8], 47% female, 56% residing in low-income zip codes) and 112,210 SAH patients (mean age [SD]: 60.2 [15.4], 62% female, 55% residing in low-income zip codes) were included (Table S1). Among the ICH cohort, 259,535 patients (mean age [SD]: 68 [14.8], 47% female, 55% residing in low-income zip codes) were admitted during the pre-pandemic period and 50,430 patients (mean age [SD]: 67 [14.9], 46% female, 57% residing in low-income zip codes) were admitted during the pandemic period. Among the SAH cohort, 93,855 patients (mean age [SD]: 60.2 [15.4], 62% female, 56% residing in low-income zip codes) were admitted during the pre-pandemic period and 18,355 patients (mean age [SD]: 60.1 [15.3], 60% female, 57% residing in low-income zip codes) were admitted during the pandemic period. In univariate analyses, ICH patients admitted during the pandemic period were significantly more likely to be insured via Medicaid (OR, 95% CI: 1.23, 1.14 – 1.33) or private (1.08, 1.01 – 1.15) insurance (vs. Medicare), have heart failure (1.13, 1.06 – 1.20), obesity (1.30, 1.21 – 1.39), renal failure (1.07, 1.01 – 1.13) and higher Charlson Comorbidity Index (1.03, 1.02 – 1.04) (Table S1).

In the pre-pandemic period, the mortality rate among ICH patients was decreasing by approximately 1% per month (aOR, 95% CI: 0.99, 0.99 – 1.00; p-value < 0.001). However, the overall mortality rate during the pandemic period increased by about 2% per month relative to the monthly rate in the pre-pandemic period (1.02, 1.00 – 1.02, p-value < 0.05) (See figure 1A and Table 1). Among patients residing in low-income zip codes, mortality rate during the pandemic period increased by 4% per month relative to the pre-pandemic period (1.04, 1.01 – 1.07). However, there was no significant change in mortality trend during the pandemic period among ICH patients residing in high-income zip codes (1.00, 0.97 – 1.03) (Table 1 and Figure 1B and 1C). Also, there was no significant change in the trend for other ICH outcomes or any SAH outcomes during the pandemic period (vs. pre-pandemic).

**Table 1:** Effect of the COVID-19 Pandemic on ICH and SAH Mortality

| **Adjusted Models** |  |  |
| --- | --- | --- |
|  | ICH Mortality | SAH Mortality |
| **Overall** |  |  |
| Pre-Covid Slope | 0.99 (0.99 – 1.00) *** | 1.00 (0.99 – 1.00) * |
| Post-Covid Slope | 1.02 (1.00 – 1.04) | 0.99 (0.96 – 1.02) |
| Difference between Pre- and Post-Covid Slope | 1.02 (1.00 – 1.04) * | 0.99 (0.96 – 1.03) |
| **Low Income** |  |  |
| Pre-Covid Slope | 1.00 (0.99 – 1.00) *** | 1.00 (0.99 – 1.00) |
| Post-Covid Slope | 1.04 (1.01 – 1.06) ** | 0.99 (0.95 – 1.03) |
| Difference between Pre- and Post-Covid Slope | 1.04 (1.01 – 1.07) ** | 0.99 (0.95 – 1.04) |
| **High Income** |  |  |
| Pre-Covid Slope | 0.99 (0.99 – 1.00) *** | 1.00 (0.99 – 1.00) |
| Post-Covid Slope | 0.99 (0.96 – 1.02) | 0.98 (0.93 – 1.03) |
| Difference between Pre- and Post-Covid Slope | 1.00 (0.97 – 1.03) | 0.99 (0.94 – 1.04) |
| **Unadjusted Models** |  |  |
|  | ICH Mortality | ICH Mortality |
| **Overall** |  |  |
| Pre-Covid Slope | 1.00 (1.00 – 1.00) *** | 1.00 (0.99 – 1.00) * |
| Post-Covid Slope | 1.01 (1.00 – 1.03) | 1.00 (0.98 – 1.03) |
| Difference between Pre- and Post-Covid Slope | 1.02 (1.00 – 1.03) | 1.01 (0.98 – 1.03) |
| **Low Income** |  |  |
| Pre-Covid Slope | 1.00 (0.99 – 1.00) ** | 1.00 (0.99 – 1.00) |
| Post-Covid Slope | 1.02 (1.00 – 1.05) * | 1.00 (0.97 – 1.04) |
| Difference between Pre- and Post-Covid Slope | 1.03 (1.01 – 1.05) * | 1.01 (0.97 – 1.04) |
| **High Income** |  |  |
| Pre-Covid Slope | 1.00 (0.99 – 1.00) ** | 1.00 (0.99 – 1.00) |
| Post-Covid Slope | 1.00 (0.97 – 1.02) | 0.99 (0.95 – 1.04) |
| Difference between Pre- and Post-Covid Slope | 1.00 (0.98 – 1.02) | 1.00 (0.96 – 1.04) |

\* - p-value < 0.05; \*\* - p-value < 0.01; \*\*\* - p-value < 0.001

**Figure 1:**
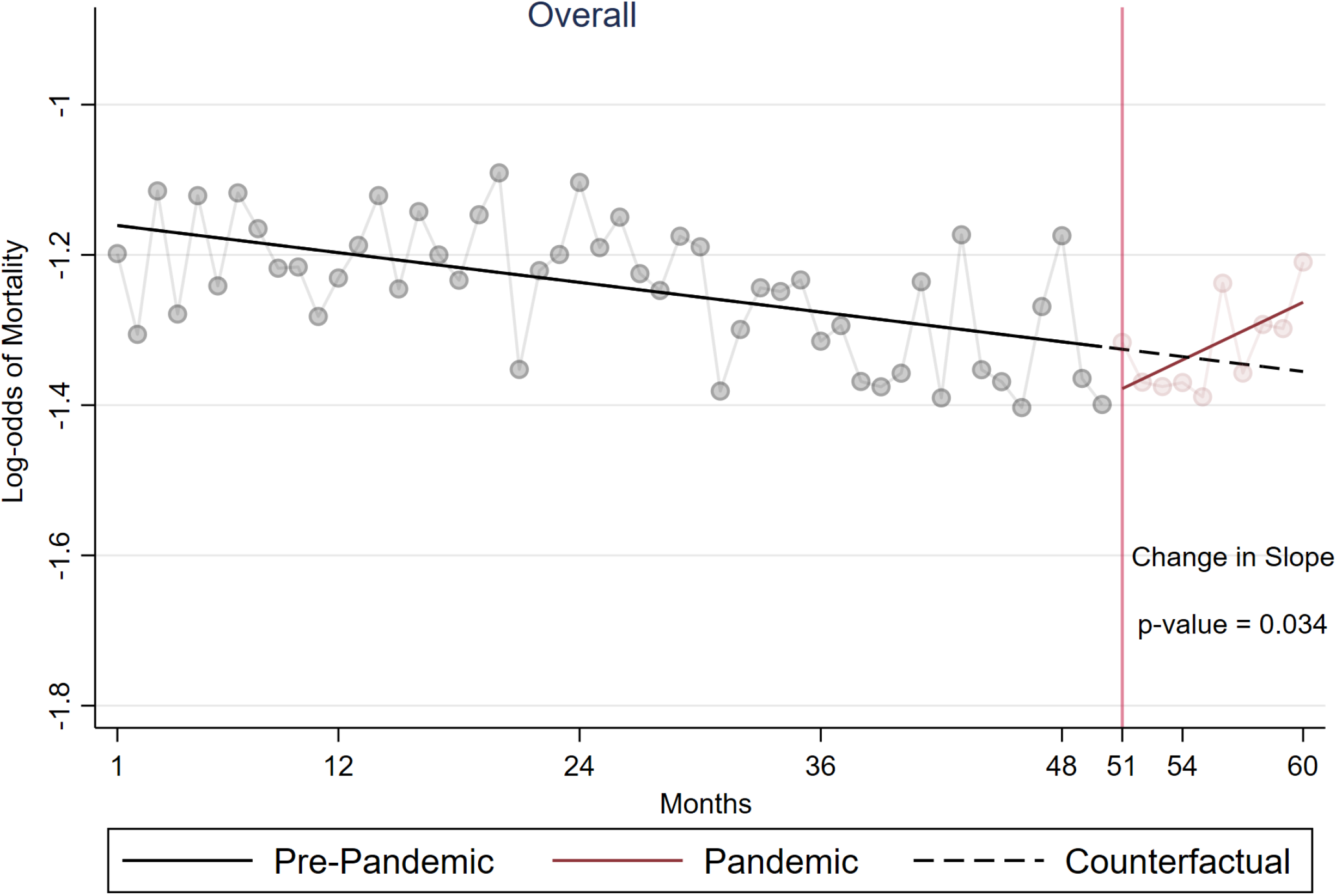
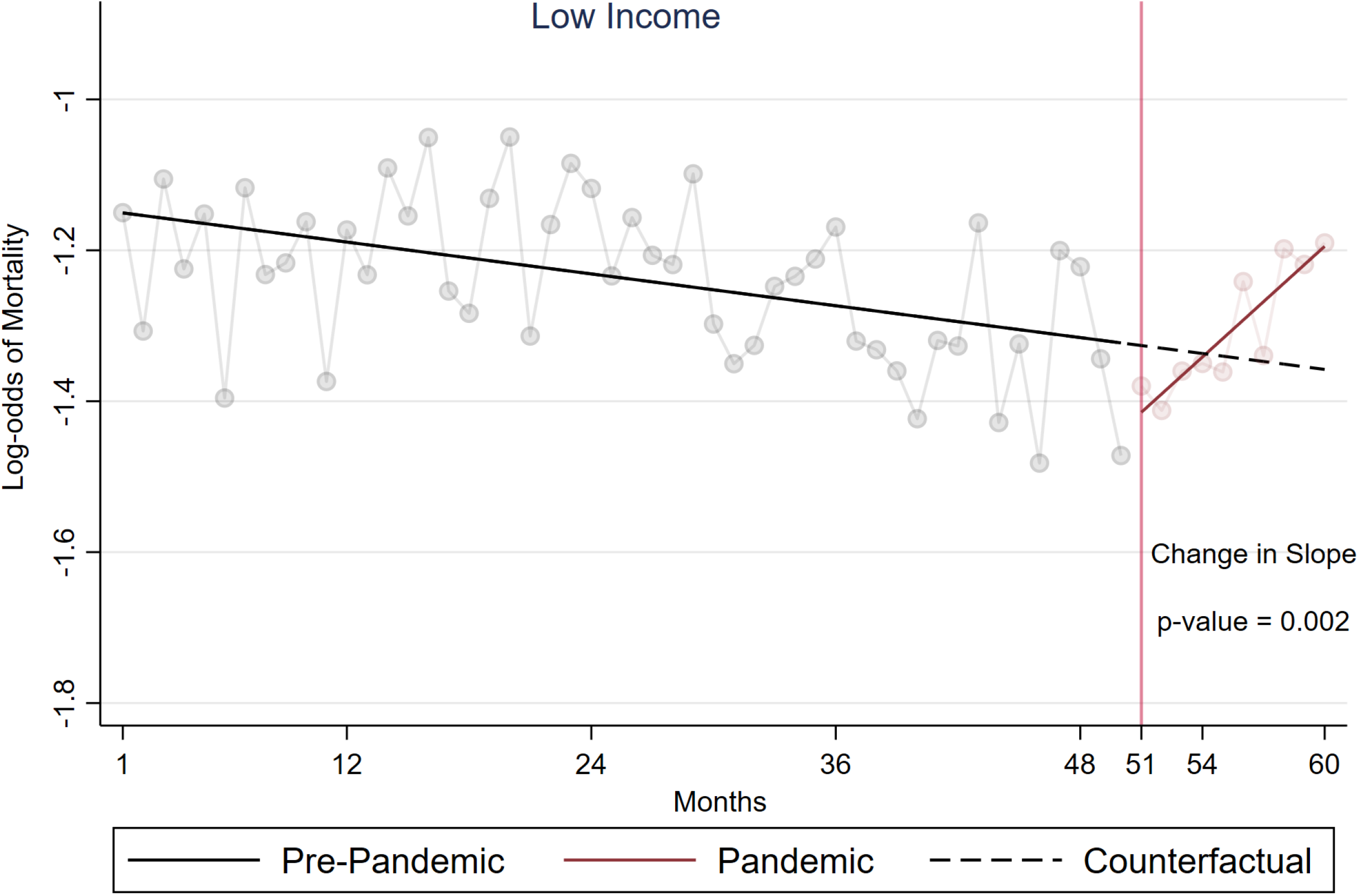
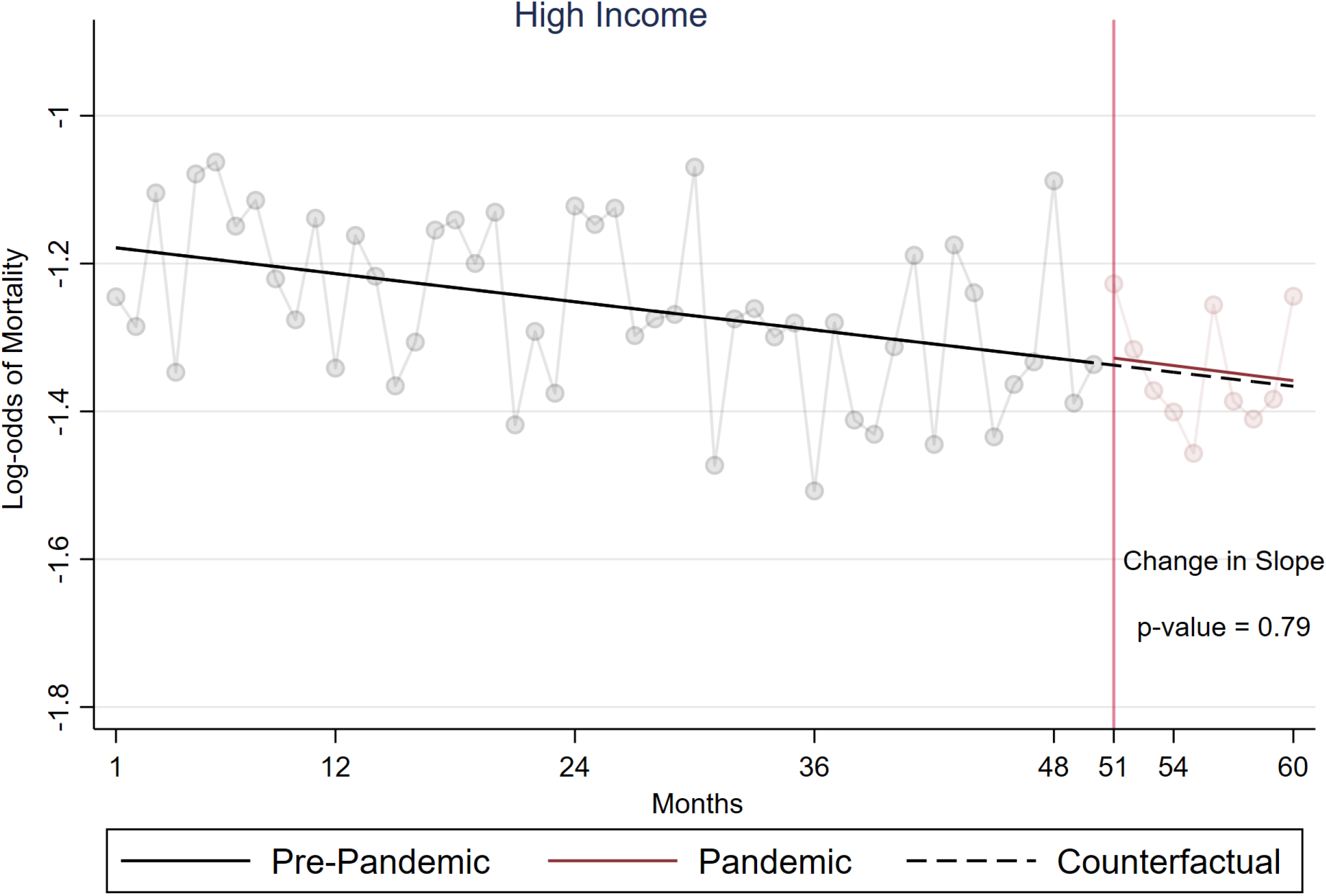
Segmented logistic regression of the effect of the COVID-19 pandemic on Intracerebral Hemorrhage Mortality, Overall (A) and disaggregated by low (B) and high (C) income residence status. Segmented logistic regression of the effect of the COVID-19 pandemic on Intracerebral Hemorrhage Mortality — unadjusted. The solid lines run through pre-intervention and post-intervention unexponentiated coefficients (logit), while the dotted lines represent what the post-pandemic trend would have been had the pandemic not occurred (counterfactual). The coefficients used for this plot have not been adjusted for confounding variables; however, the reported p-values for the difference in slope between pre-pandemic and post-pandemic period have been adjusted for confounding. A p-value < 0.05 indicates that there is a significant change in trend (slope) between the pre-pandemic and post-pandemic mortality rate.

Among patients admitted between April and December 2020 (ICH: 44,405 without COVID-19 and 935 with COVID-19; SAH: 16,205 without COVID-19 and 395 with COVID-19), males (aOR, 95% CI: 1.42, 1.03 – 1.97) (vs. females), NHB (1.94, 1.28 – 2.95), Hispanics (3.59, 2.29 – 5.64) and the “Other” race/ethnicity category (3.66, 2.16 – 6.19) (vs. NHW) had significantly higher odds of having comorbid ICH and COVID-19, while Hispanics (vs. NHW) have significantly higher odds of having comorbid SAH and COVID-19 (4.73, 2.88 – 7.79) (Figure 2).

**Figure 2:**
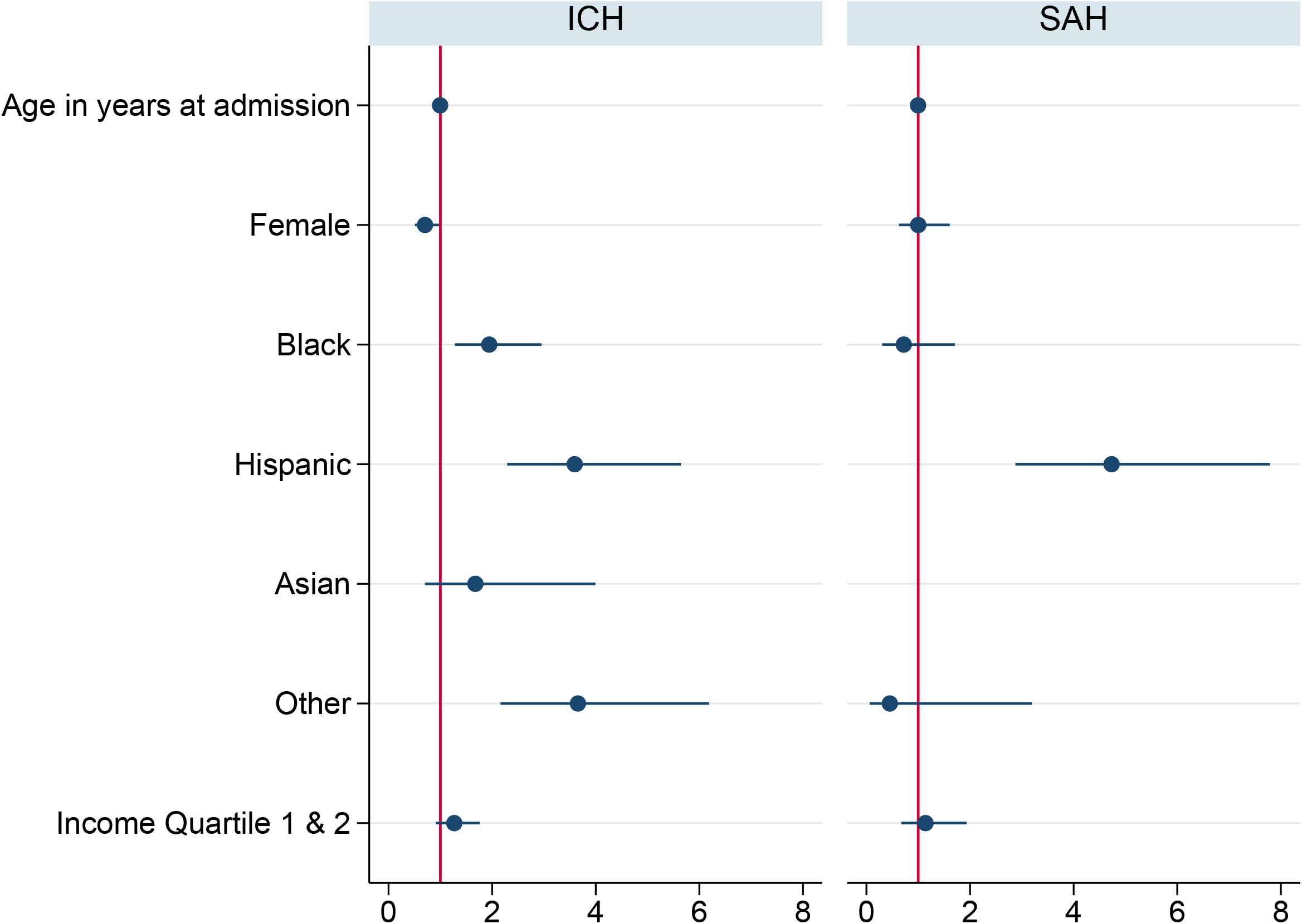
Sociodemographic factors associated with having comorbid COVID-19 and ICH/SAH.

In multivariable analyses, ICH and SAH patients with comorbid COVID-19 had a significantly higher likelihood of mortality compared to patients admitted between April to December 2019, overall (aOR, 95% CI: 1.83, 1.33 – 2.51 for ICH; and 2.76, 1.68 – 4.54 for SAH) and among patients with MM-LoF (2.15, 1.12 – 4.16 for ICH; and 5.77, 1.57 – 21.17 for SAH). However, among patients with E-LoF, there was no significant difference in the likelihood of mortality between ICH and SAH patients with comorbid COVID-19 admitted between April to December 2020 and patients admitted during a similar period in 2019. Furthermore, among ICH and SAH patients without comorbid COVID-19, the likelihood of mortality was similar across the 2020 and 2019 April to December cohorts.

Tables S2 and S3 provide details of the univariate comparisons of the characteristics of ICH and SAH patients with comorbid COVID-19 (admitted between April to December 2020) and patients admitted during a similar period in 2019 (model group 1). Tables S2 and S3 also provide univariate comparisons of ICH and SAH patients admitted between April and December 2020 with and without comorbid COVID-19 (model group 2).

## Discussion

We evaluated the association of the COVID-19 pandemic with ICH and SAH in-hospital outcomes in a nationally representative sample. Relative to the pre-pandemic period, we observed a significant increase in the monthly rate of in-hospital mortality among ICH patients during the pandemic period. This increase was primarily driven by ICH patients residing in low-income zip codes, whereas no change in mortality was observed among patients residing in high-income zip codes. We also demonstrate that comorbid COVID-19 was associated with higher likelihood of mortality among ICH and SAH patients with MM-LoF, but not among patients with E-LoF.

Similar to a previous report,^12^ our analyses demonstrate that ICH mortality was significantly declining during the pre-pandemic period. However, this trend was reversed during the pandemic period, particularly among patients residing in low-income zip codes. Relative to the pre-pandemic period, the overall ICH mortality rate increased by 2% per month in the pandemic period. This acceleration of mortality rate seems to be largely driven by patients residing in low-income zip codes, among whom ICH mortality rate increased by 4% per month during the pandemic period, whereas no significant change in mortality was observed among patients residing in high-income zip codes during the pandemic period. These findings suggest that the COVID-19 pandemic may have slowed down the sustained improvement in ICH mortality observed during the pre-pandemic period, particularly among the low-income population. Though our analyses do not definitively outline the reasons for disparate COVID-19 associated ICH outcomes; higher comorbidity burden, lack of access, awareness, and even disparities in care (including delayed care) may be postulated as potential drivers of such disparities. Most importantly, our analyses are yet another demonstration of the pandemic’s disproportionate impact on vulnerable populations and highlight the need for continued focus on uncovering and addressing the reasons for the now widely reported socioeconomic disparities, particularly among patients with cerebrovascular disease.^13^

Similar to prior smaller studies, we also report that hemorrhagic stroke (ICH and SAH) patients with comorbid COVID-19 have significantly higher mortality compared to patients without COVID-19.^14,15^ However, our data uniquely demonstrates, at the national level, that comorbid COVID-19 was only associated with a higher likelihood of in-hospital mortality among ICH and SAH patients with MM-LoF, whereas among patients with E-LoF, COVID-19 status was not a significant driver of mortality. These findings have significant clinical relevance, and though we are limited from conducting a clinically detailed exploration of the biological mechanisms driving the differences in mortality between hemorrhagic stroke patients with and without comorbid COVID-19, it is reasonable to surmise, from previous studies, that a heightened systematic inflammatory response to the COVID-19 virus and it’s directed end organ damage may be potentiating these poor outcomes.^16^ However, further studies are needed to elucidate the mechanisms driving poorer outcomes among hemorrhagic stroke patients with comorbid COVID-19. Also, given that minority races/ethnicities are at a higher likelihood of having comorbid COVID-19, the findings of this research highlight the need to further investigate the biological and environmental factors potentially driving socioeconomic disparities in the association between COVID-19 and hemorrhagic stroke outcomes.

Our study has some limitations. First, this study covers only the first wave of the COVID-19 pandemic. Hence, future studies are needed to explore the trends in the subsequent waves of the pandemic as data for ensuing years become available. Secondly, our analysis may have missed COVID-19 patients who did not require or receive in-hospital care for COVID-19 and potentially underestimated the prevalence of COVID-19 among patients with ICH and SAH. Thirdly, we did not have detailed data on the timing of hemorrhagic stroke and COVID-19 diagnosis. Finally, we did not have access to more granular clinical data, including patients’ imaging data (to ascertain hemorrhage location, volume, or other hemorrhage characteristics) and information on the COVID-19 variants. Nevertheless, the insights provided by this study will be useful in guiding the readiness of public health authorities to implement strategies addressing sociodemographic disparities during public health emergencies.

## Conclusions

The study found a significant acceleration of in-hospital mortality rate among ICH patients during the post-pandemic period, particularly among those residing in low-income zip codes. Sustained efforts are needed to better understand the impact of the pandemic on stroke outcomes, particularly among vulnerable populations, as well as to address disparities in healthcare access, quality, and outcomes during public health emergencies.

## Data Availability

After completing a data use agreement training, qualified researchers can obtain NIS data through the Health Care Utilization Project?s central distributor

https://www.distributor.hcup-us.ahrq.gov/

## Acknowledgments

Dr Bako had full access to all the data in the study and takes responsibility for the integrity of the data and the accuracy of the data analysis. The content of this article is based on data from the National Inpatient Sample (NIS) database, acquired from the Agency for Healthcare Research and Quality’s (AHRQ) Health Care Utilization Project (HCUP). However, the content of this article does not necessarily reflect the views and opinions of the AHRQ. Also, the AHRQ did not participate in the study’s design, implementation, data analysis, interpretation, nor manuscript preparation, review, approval, or decision to submit the article for publication.

